# The unmutated common ancestor of Gp41 conformational epitope targeting variable heavy chain 1-02 utilizing antibodies maintains antibody dependent cell cytotoxicity

**DOI:** 10.1101/2021.09.23.21264029

**Authors:** Brian H. Wrotniak, Meghan Garrett, Sarah Baron, Hakimuddin Sojar, Alyssa Shon, Elizabeth Asiago-Reddy, Jessica Yager, Spyros Kalams, Michael Croix, Mark D Hicar

## Abstract

In studies on monoclonal Abs (mAbs) from long-term non-progressors (LTNPs), our laboratory has previously described highly mutated Abs against a complex conformational epitope with contributions from both gp41 heptad repeat regions. Despite using the VH1-02 gene segment, known to contribute to some of the broadest neutralizing Abs against HIV, members of these Abs, termed group 76C Abs, did not exhibit broad neutralization.

Because of the excessive mutations and use of VH1-02, our goal was to characterize the non-neutralizing functions of Abs of group 76C, to assess targeting of the epitope in various clinical presentations, and to assess the development of these Abs by comparison to their predicted common ancestor. Serum competition assays showed group 76C Abs were enriched in LTNPs, in comparison to VRC-01. Specific group 76C clones 6F5 and 6F11, expressed as recombinant Abs, both have robust ADCC activity, despite their sequence disparity. Sequence analysis predicted the common ancestor of this clonal group would utilize the germline non-mutated variable gene. We produced a recombinant ancestor Ab (76Canc) with a heavy chain utilizing the germline variable gene sequence paired to the 6F5 light chain. Competition with group 76C recombinant Ab 6F5 confirms 76Canc binds HIV envelope constructs near the original group C epitope. 76Canc demonstrates comparable ADCC to 6F5 and 6F11 when targeting both clade B and C HIV constructs. The functional capability of Abs utilizing germline VH1-02 has implications for disease control and vaccine development.

## Introduction

The creation of a successful HIV vaccine continues to be a public health priority. A large effort has been focused on discovery and characterization of broadly neutralizing Abs (bnAbs). Whether similar bnAbs can be induced from stimulation of native B cells during immunization has been a challenge in HIV vaccine design. We previously used HIV virus-like particles to bind B cells from long-term non-progressor (LTNP) subjects. From these B cells, a number of mAbs were cloned and showed an extreme number of mutations, similar to many of the known bnAbs. When expressed as recombinant full-length Abs, some showed specificity or higher affinity for trimerized envelope and virus-like particles, which we termed quaternary-targeting Abs (QtAbs) (Hicar, Chen, Sulli, et al., 2016). Despite the highly mutated nature of these Abs, they are generally non-neutralizing Abs (nnAbs) (Hicar, Chen, Kalams, et al., 2016).

Recent studies have shown that a number of these highly mutated QtAbs have the capacity to mediate Ab dependent cell cytotoxicity (ADCC) (Sojar et al., 2019). ADCC was implicated in the RV144 vaccine trial success (Haynes et al., 2012). The Fc portion of Abs, which ADCC is dependent on, is crucial in protection (Hessell et al., 2007). Passive Ab infusions of both neutralizing (Hessell et al., 2009; Parren et al., 1995; Parren et al., 1996) and non-neutralizing Abs protect macaques in a SHIV mucosal challenge model (Santra et al., 2015). NnAbs targeting structural epitopes in gp41 have shown ADCC function and limiting of cell-to-cell spread of infection (Bonsignori et al., 2012). Other nnAbs targeting this same gp41 epitope, such as F240 (Burton et al., 2011) and 7B2 (Santra et al., 2015), can protect macaques during SHIV vaginal challenge. ADCC-mediating Abs from an HIV-1 vaccine efficacy trial target multiple epitopes and preferentially use the VH1 gene family (Bonsignori et al., 2012). ADCC activity of Abs using VH1 genes correlated with the degree of somatic mutation. ADCC activity has been associated with control of HIV in the past (Lambotte et al., 2009) and more recent polyfunctionality, including ADCC, of Abs has been shown to correlate with control (Das et al., 2020).

We have previously mapped another highly mutated (83% homologous to predicted heavy chain germline) QtAb, mAb 76-Q13-6F5 (6F5), to a novel structural epitope. The 6F5 epitope encompasses areas in both heptad repeats of gp41, mapping by alanine scanning mutagenesis to amino acids R557, E654 and E657 of reference sequence HXB2 (Hicar, Chen, Sulli, et al., 2016). Since alanine scanning mutagenesis showed widely separated amino acids as potentially involved, this suggested a structural epitope that is available on post-fusion forms. From these studies, Abs from other genetically related groups (Q7 and Q11) also showed that E657 was critical for binding using alanine scanning mutagenesis. On sequence analysis, 10076-Q7-6F11 (6F11), 10076-Q7-7C6 (7C6), 10076-Q11-4E4 (4E4) and 6F5 had the same CDR3 length and, the same predicted germline sequences for both heavy (VH1-02) and light chains (VK1-39), but had < 90% of shared nucleotide sequence (Hicar, Chen, Sulli, et al., 2016). 6F11, 7C6, and 4E4 also competed with 6F5 binding to HIV envelope trimers, so these were classified into a single epitope group (group 76C). Despite the low homology, these similarities and the similar epitope mapping implied these were once distantly related to a single B cell lineage.

Since group 76C Abs bind trimers and interfere with the function of fusion inhibitors (Smith et al., 2018), the targeted epitope is at least partially available in the pre-fusion state. Since fusion of the viral membrane to the target cell depends on heptad repeat regions associating and forming a six-helix-bundle (6HB) post-fusion, these group 76C Abs allow us to explore development of functional targeting to this complex gp41 epitope. Because of the wide disparity in the mature group 76C Abs, the predicted common ancestor of these Abs would utilize germline VH1-02. From exploring the derivation and possible cross-reactivity of such Abs, we discovered that the germline VH1-02 gene can support ADCC functional activity.

## Materials & Methods

### Clinical Samples

LTNP were defined as those individuals who maintained CD4 counts > 300/mm^3^ for minimally 7 years without therapy post-diagnosis. LTNP samples were collected from State University of New York clinical sites and shared as codified samples with associated clinical information under the UB IRB STUDY00000359. Biobanked samples from controls and additional LTNP samples accessed through the Biorepository at the University of Washington, through collaboration with receipt of deidentified samples from Vanderbilt University. Controls requested from the biorepository were infected for over 7 years and did not continually maintain CD4s and/or went on therapy prior to the seventh year of infection.

### Statistical analysis

For the serum comparison of LTNP to other HIV infected, power calculations for 80% power with alpha of 0.05 based on means of 50% of inhibition versus 60% inhibition with relatively wide variance (10%) suggested we would need minimally 16 members in each group. The levels of binding antibodies in LTNPs were compared to serum from a group of HIV infected controls with comparable viral loads using Mann–Whitney U tests (two-tailed and alpha set at 0.05). Analyses were conducted using SYSTAT software version 13 (SYSTAT Software, 2004), with statistical tests applied as described in the figure legends. Graphpad Prism 9 was used for graphical representation of data was, with means and standard deviations shown unless otherwise noted.

### 76 Group C antibodies

During the initial characterization of recombinant Ab binding, the mAb 6F5 competed with three other Abs:76-Q7-6F11 (6F11), 76-Q7-7C6 (7C6), and 76-Q11-4E4 (4E4) (Hicar, Chen, Sulli, et al., 2016). These Abs shared the same predicted variable chain sequences (VH1-2, VK1-39) and had the same CDR lengths, consistent with a distal clonal relationship. Their original clone names contain the subject (76 for 10076),-Qxx(for quaternary targeting clonal group), and then the specific Ab reference. These were originally assigned to different groups (Q7, Q11, and Q13) using the definition of clonality of having same heavy chain CDR3 region length, >90% CDR3 nucleotide identity, and having overall amino acid sequence homology of > 90%. Two other heavy chains (76-Q2-1F7 and 76-Q2-1B1) had similar variable gene usage and structure, but did not have light chain pairs isolated so were not characterized as recombinant Abs. Within this manuscript, these Abs are referred to by their shortened specific clonal names (*i*.*e*. 6F5) (Hicar, Chen, Kalams, et al., 2016).

### Full-length Ab expression

After cloning into an expression vector, heavy and light chain Ab plasmid DNA were amplified using the Pure Yield Plasmid Maxiprep System (Promega #A2392) following the manufacturer ‘s instructions. Plasmids were then co-transfected into Free Style 293-F cells. Briefly, 50 μg of DNA per 100 ml of culture was mixed with 3 ml of Opti-MEM I Reduced Serum Medium (Thermo Scientific #31985062) and sterile filtered. Next, 100 ul of 1mg/ml polyethylenimine (PEI) per 100 ml of culture was added to 3 ml of Opti-MEM I reduced Serum Medium and sterile filtered. The DNA and PEI solutions were then mixed dropwise, vortexed, and incubated for 12 minutes at room temperature. The mixture was added to 100 ml of Free Style 293-F cells seeded in Free Style 293 Expression Medium (Thermo Scientific #12338018) at a density of 1×10^6^ cells/ml and left to incubate for six days at 37°C 8% CO2 on a shaker with 130 rpm speed. On the 6^th^ day of transfection, culture supernatants were collected and cleared by centrifugation at 3000 x *g* for 15 minutes at 4°C and then filtered through a 0.2 μm filter. Following filtration, immunoglobulins were purified using a 5 ml HiTrap Protein G column (GE Healthcare 17-0404-01) on an AKTA FPLC (GE Healthcare, Piscataway NJ). The IgG fraction was recovered from the column by elution with glycine buffer (100 mM, pH 2.5). The fractions containing IgG were pooled and exchanged with 1x PBS (pH 7.4) via 3 washes using Amicon Ultra-2mL Ultracel-10kDa cutoff (Millipore Corp., Bedford, MA). A Tris-HCl neutralization step was avoided as it may interfere with later biotinylation steps. Ab concentrations were measured on a Nano Drop Lite Spectrophotometer (Thermo Scientific).

### Ab binding ELISAs

Immulon 2-HB plates were coated with respective antigen in 0.1 mM sodium bicarbonate buffer pH 8.4 overnight at 4°C.The next day plates were blocked with 10% Fetal bovine serum (FBS) in PBS for 2 hrs at 4°C. After removing blocking buffer, 100 ul of respective Ab diluted in 10% FBS was added and incubated at 37°C for 1 hr. After 1 hr ,plates were washed with washing buffer (PBS containing 0.1% Tween 20) three times. After washing 100 ul of HRP conjugated secondary Ab in blocking buffer was added and incubated at room temperature for 1 hr. Plates were washed three times with washing buffer and 50ul Ultra-TMB (Thermo Scientific) substrate was added to each well and the reaction was stopped by adding 50ul of 2N sulfuric acid. Absorbance was measured at 450nM optical density within 30 min.

### Ab biotinylation competition ELISAs

Ab biotinylation was performed as previously described (Hicar, Chen, Sulli, et al., 2016). First, 100-200 μl of 3 mg/ml Abs were equilibrated in carbonate buffer (pH 9.3) using a 10k or 50k cutoff Amicon Ultra-4 Centrifugal Filter Unit (Millipore Corp., Billerica, MA) per the manufacturer ‘s instructions. Ab concentrations were measured using a Nanodrop spectrophotometer (Thermo Scientific, Grand Island, NY). Sulfosuccinimide biotin (Sigma-Aldrich, St. Louis, MO) was added at a 30-fold molar excess over the concentration of Ab (approximately 90 μg per mg of Ab). This solution was mixed and incubated for 30 minutes with gentle shaking at room temperature. Tris-HCl (pH 7.5) was added up to 10% by volume to bind residual biotin molecules. The biotinylated Ab was separated from residual biotin complexes and equilibrated with PBS (pH 7.4) using a G-25 Sephadex Nap-5 (GE Healthcare, Piscataway NJ) column per the manufacturer ‘s protocol. Biotinylated Abs were competed with a panel of known unlabeled HIV-specific Abs, as listed in the results section. ELISAs were performed similar to previous description, with streptavidin-HRP (Southern Biotech, Birmingham, Al) conjugated secondary Ab reagent for detection of binding.

### Western blot analysis

After running on SDS-PAGE, proteins were transferred onto blotting membrane using Iblot dry blotting system (Thermo Fisher Scientific) according to the manufacturer ‘s instructions. After transfer, the blot was blocked for 1 hr at room temperature. After rinsing, primary Ab was diluted in 10% FBS was added and incubated overnight at 4°C. Blot was washed (3 × 10 minutes) with gentle agitation. Secondary Ab (Alkaline phosphatase - conjugated anti-human IgG, Southern Biotech, Birmingham, AL) was added in wash buffer and incubated for 1 hour at room temperature with gentle agitation. Blot was then washed 5×5minutes, three times in PBST and twice in PBS. Bands were visualized with Alkaline phosphate substrate NBT/BCIP (Thermo Scientific, Grand Island, NY).

### Six-helix-bundle (6HB) formation and binding

Protocol followed previous publication (Liu, Zhao, & Jiang, 2003). Peptide N36 in PBS was incubated with C34 at 37°C for 1 hr. The mixture was added to 96-well plates which were precoated with NC-1 IgG (1ug/ml) purified from mouse antisera directed against the gp41 6HB. The plate was incubated for 2 hr at room temperature. Plate was extensively washed with washing buffer containing Tween-20. Then primary Abs were added at various dilutions and incubated at 37°C for 1 hr. The plate was washed extensively with washing buffer and secondary HRP-conjugated anti human IgG (H+L) at 1:2000 dilution was added in each well and incubated at room temperature for 1 hr. The plate was washed and TMB substrate was added for color development and the reaction was stopped using 2N sulfuric acid. Human anti-HIV-1 gp41 Ab HK20 was used as a control (Creative Biolabs, Shirley, NY).

### Ab dependent cellular cytotoxicity assay

Rapid and fluorometric ADCC (RFADCC) assay was performed as described previously (Milligan, Richardson, John-Stewart, Nduati, & Overbaugh, 2015; Williams et al., 2015). Briefly, CEM-NKr target cells were double stained with PKH (a membrane stain) and CFSE (a cytosolic stain) and coated with 50ug/ml ZA 1197 MNgp41 (NIH AIDS Reagent Program). The coated target cells (5000 cells/well) were incubated with the Ab for 10 minutes to allow binding of the Ab to gp41. Next, effector cells (HIV-negative donor PBMCs) were added at an effector target ratio of 50:1. The mixture was incubated for 4 hours at 37^0^ C. After the incubation, the cells were fixed in 1% paraformaldehyde in PBS. When ADCC occurred, the target cell membrane is permeabilized leading to the loss of CFSE and retention of PKH. ADCC was measured by flow cytometry as the percentage of PE (+) CFSE (-) cells among all PE (+) cells with background (average ADCC mediated against uncoated target cells) subtracted. Background was set to 3-5%. Influenza Ab F16V3 (500 ng/uL) was included as a negative control and results are normalized to HIV immunoglobulin (HIVIG, 500 ng/uL).

## Results

### Enrichment for Antibodies binding against 76C epitope in Long-term non-progressors

Recent studies support that specific polyfunctional Abs may influence disease progression (Das et al., 2020). Prior studies that showed that ADCC was globally higher in controllers, also studied specific epitope targeting by serum competition with biotinylated Abs. No differences in Abs targeting the CD4 binding site or other epitopes associated with neutralization were shown between viremic individuals and viral controllers (Lambotte et al., 2009). The 76C group Abs target a previously unassessed conformational epitope in gp41. We used 6F11 as the representative biotinylated Ab since it had a higher affinity for HIV BaL foldon trimers (Hicar, Chen, Kalams, et al., 2016), the target we chose for this assay. Samples were obtained from recruitment and biobank access from 35 LTNP individuals (Minimally 7 years off therapy with stable CD4 counts), and 58 HIV infected individuals with comparable viral loads. We compared the pattern of Ab targeting in these groups between a 76C Ab (6F11) and the CD4 binding site Ab VRC01 which also utilizes VH1-02 (**Figure 1**); this serves as a control for inherent immunologic differences (CD4 helper variation, viral load associated inflammation, etc.). Abs that can compete with group 76C Abs were more prevalent in the cohort of LTNPs (p<0.01). Consistent with prior studies (Lambotte et al., 2009) the CD4 targeting Ab VRC01 showed no difference between LTNPs and those with low CD4 cells.

**Figure 1:**
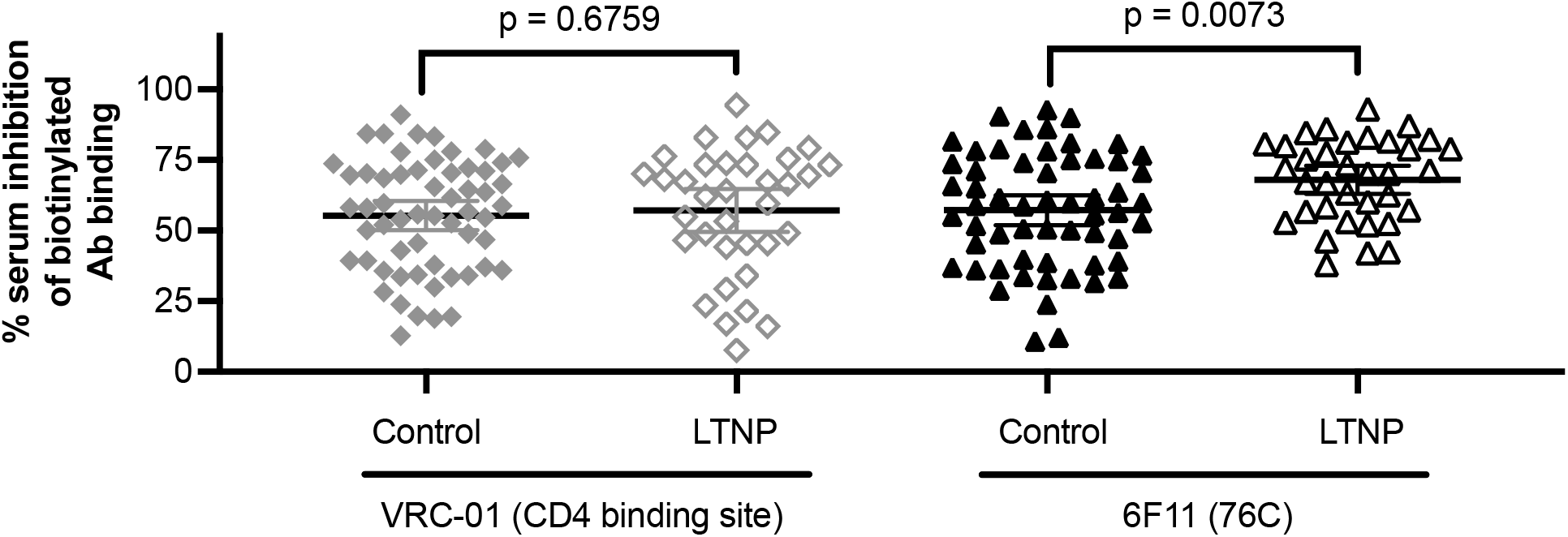
Serum from LTNPs is enriched for Abs targeting the 76C complex conformational epitope. Serum from LTNPs compared to serum from a group of HIV infected controls with comparable viral loads were used to compete against biotinylated CD4 binding site (VRC01; 1:200 serum dilution) and 76C Gp41 conformational epitope (6F11; 1:800 serum dilution) targeting Abs. Serum dilutions were chosen to align data means near 50% to avoid data compression. Data were normalized to inverse of percent biotinylated Ab binding compared to positive and negative control range. Experiments were repeated twice with representative results of single experiment shown. Means with 95% confidence intervals are shown. Differences between groups were assessed with a Mann-Whitney U test for each serum dilution.

### Functional activity

In prior studies, group 76C Abs did not neutralize various tiers of HIV strains (Hicar, Chen, Kalams, et al., 2016). We have recently shown that members of this group 76C can interfere in certain fusion interference assays implying that a portion of this epitope is available pre-fusion (Smith et al.,2018). In the original studies that identified 6F5, the clade B virus-like particle also identified the Ab 76A-2C6 (classified as group A due to targeting of a separate epitope). 76A-2C6 targets a complex conformational epitope, recognizes multiple clades, and was shown to have robust ADCC activity using both clade B and clade C constructs (Sojar et al., 2019). So, we chose to test for ADCC activity in these group C Abs using clade B MN and clade C ZA1197 gp41 constructs. Group 76C members 6F5 and 6F11 both showed comparable ADCC activity to 76A-2C6, QA255.006 and HIVIG (**Figure 2**).

**Figure 2:**
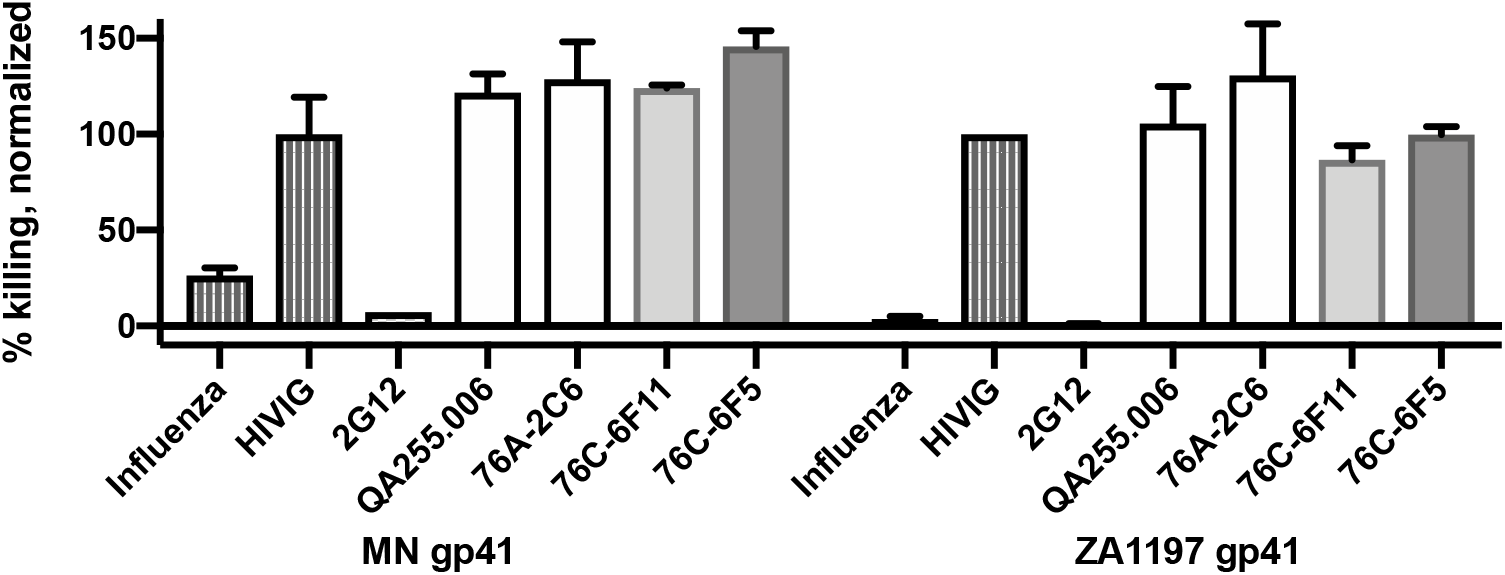
Antibody dependent cell cytotoxicity of group 76C Abs 6F5 and 6F11. Percent killing of cells coated with MN gp41 and ZA 1197 gp41 mediated by group 76C Abs 6F11 and 6F5 are shown. Results are compared to negative control bnAb 2G12, anti-influenza Ab F16V3 and positive controls HIVIG, Ab QA255.006 (gp41 Ab with ADCC), and the structural targeting Ab 76A-2C6. Results shown represent a single experiment (with two replicates) for group MN gp41 and the combined data of two experiments using two different donors for ZA1197 gp41.

### Sequence, creation of group 76C ancestor

6F5 binds a conformational epitope on gp41. On analysis for common shared mutations, besides those noted within the CDR3 regions, there were no shared mutations from germline across all six putative members in heavy chain or of the four that had cloned light chains (**Figure 3**). This surprisingly suggested a propensity for germline variable chain binding to target this antigenic site. Notably 6F11 and 7C6 had the exact same CDR3 segment in both the heavy and light chains. 4E4 shared rare mutations with all 76C members and was the least mutated (87.5% homology to predicted germline) of the 76C members. Thus, we created an ancestor Ab (76Canc) by expressing the 6F5 light chain with a heavy chain construct utilizing the germline VH1-02 sequence (through framework 3) and the heavy chain CDR3 sequence of mAb 4E4.

**Figure 3:**
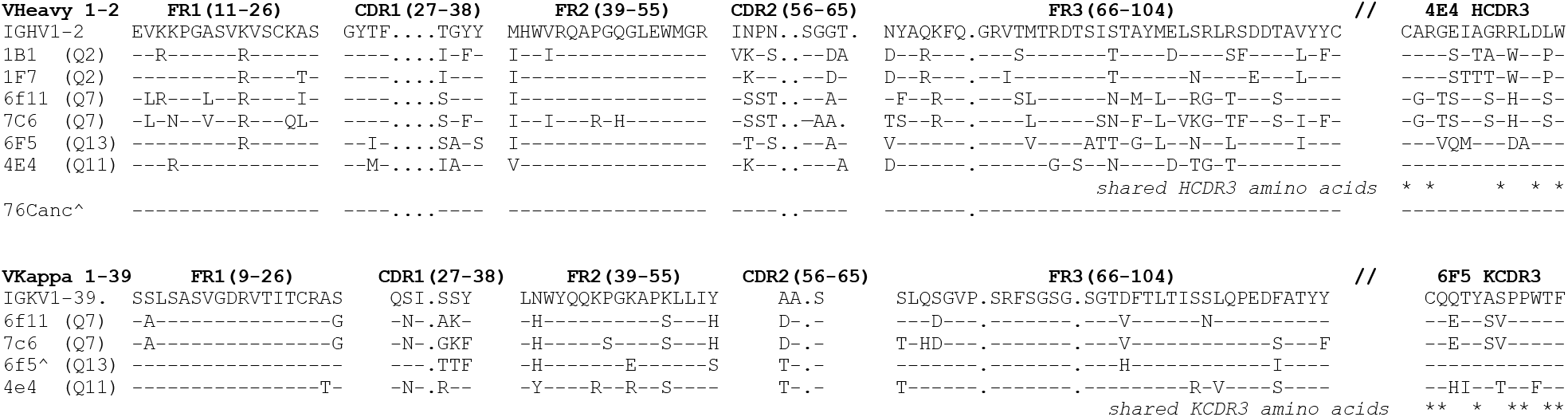
Sequence comparison of group 76C Abs supports that germline VH1-02 can contribute to epitope recognition. Comparison of the group 76C Abs (original clonal assignment by genetic relationship is shown; Q#, see methods) shows no mutations from germline shared in all clones. Despite the shared length, there are few shared amino acids in the CDR3 regions (*). Germline 4E4 HCDR3 contributed to the final 76anc construct. Gaps in IMGT numbering system are shown (.). Amino acids shared with predicted germline sequence are noted by dashes (-) and mutations away from germline are noted by their predicted single amino acid designation. ^76Canc heavy chain and 6F5 light chain was used in the recombinant 76Canc antibody.

### Ab 6F5 competition

We initially compared HIV antigen binding of 76Canc to 6F5, 76A-2C6, and negative control 8B10. 76Canc showed specific recognition of BaL trimers (**Figure 4A**) and gp41 clade B protein (**Figure 4B**), but this was less than both 6F5 and 6F11. Results from 6F11, 76A-2C6 and 6F5 had comparable EC50 estimates to those previously published (Hicar, Chen, Sulli, et al., 2016), as 76A-2C6 has a notably increased specificity for trimeric forms of envelope. From gp41 binding, the estimated EC50 of 76Canc was 113.6 (ng/ml). To confirm antigen targeting of 76Canc we performed competition ELISA using biotinylated 6F5 (6F5-biotin) recombinant Ab with Trimer (**Figure 4C**) as well as gp41 (**Figure 4D**). 76Canc partially competed with 6F5-biotin for both antigens, though more efficiently on gp41 (compared to non-competitive bindind signified by ‘blank ‘ well). Notably, both 6F11 and 6F5 showed more efficient 6F5-biotin competition.

**Figure 4:**
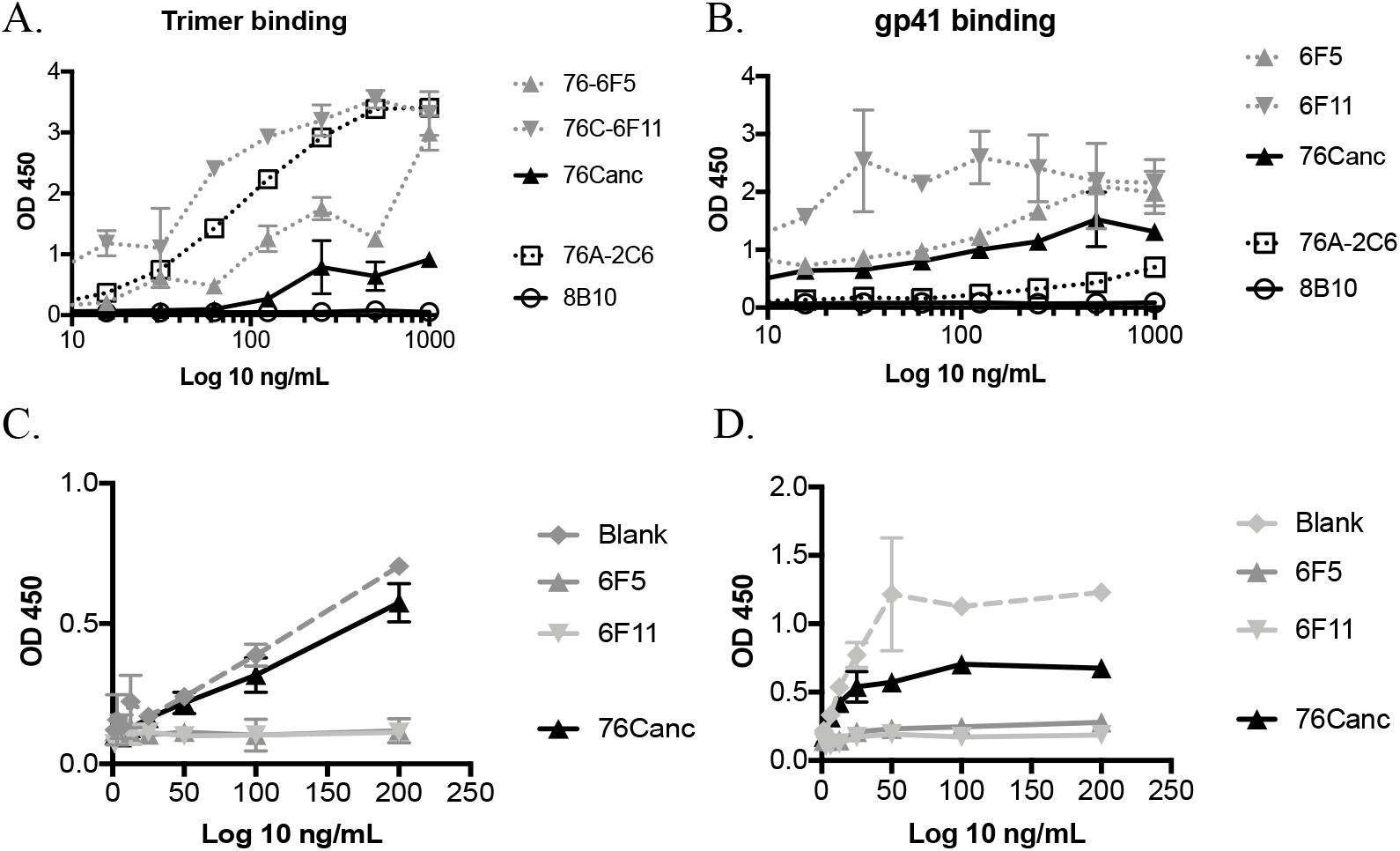
Binding of representative group 76C Abs and competition against biotinylated 6F5. ELISA binding of representative group 76C Abs to A) trimer, and B) gp41 with 76A-2C6 as positive control and 8B10 as negative control. Competition of biotinylated 6F5 antibody with group 76C Abs on C) trimer and D) gp41. Blank signifies no competitive Ab addition. Experiments were repeated thrice with representative results of single experiment shown.

### Post fusion six helix bundle (6HB) binding

Our earlier study (Hicar, Chen, Sulli, et al., 2016) showed mutations widely separated on the primary sequence (R557, E654 and E657; numbering per HXB2 reference sequence) affected 6F5 binding. To confirm these Abs can target the post-fusion forms of gp41, we used peptides previously described that can replicate this post-fusion form. The two peptides N36 (SGIVQQQNNLLRAIEAQQHLLQLTVWGIKQLQARI) and C34 (WMEWDREINNYTSLIHSLIEESQNQQEKNEQELL), each containing one of the heptad repeats, can form the post-fusion 6HB in vitro, which can be visualized by gel shift (Liu et al., C. 2003). On native gel PAGE (**Figure 5A**), the N36 peptide is not shown as it ‘s higher pI of 11.2, prevented it from entering the gel. With mixing prior to gel loading of the C34 peptide (acidic pI of 3.4) with N36, an appreciable shift is noted compared to C34 lane alone (**Figure 5A**). The 6HB formation was confirmed by probing with NC-1 Ab (**Figure 5B**); an Ab specific to the post-fusion form as shown by lack of binding to C34 alone (Jiang, Lin, & Lu). On sandwich ELISA testing using NC-1 as a capture Ab, 6F11, 6F5, and 76Canc all bound the 6HB comparable to HK20, an Ab that targets the 6HB. Further, a 6HB specific interaction was confirmed using western blot and decaprobe by running 6HB preparative gel on charge basis (**Figure 5D**). 6HB binding of these group 76 Abs was further confirmed by competition with 6F5-biotin (**Figure 5E**). Again, the 76Canc partially competes with 6F5 binding, where 6F11 and 6F5 more efficiently compete.

**Figure 5:**
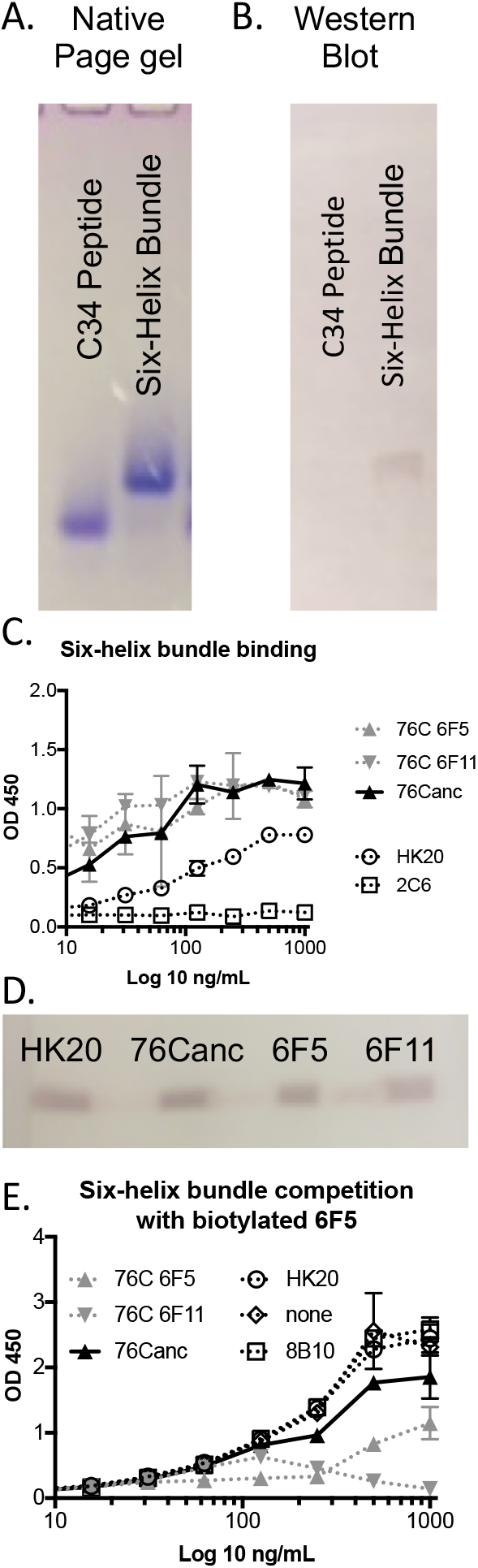
Six-helix-bundle (6HB) formation and binding of various Abs. A) Coomassie blue staining of C34 & 6HB formed by incubating N36 and C34. B) Western blot of C34 & 6HB with anti-mouse NC1 monoclonal Ab. C) 6HB sandwich ELISA binding using capture Ab NC1. D) Western blot analysis of 6HB binding of various Abs using decaprobe. E) 6HB binding competition against biotinylated 6F5 Ab. Experiments were repeated at least twice with representative results from single experiments shown.

### Functional activity of Germline based 76Canc

Because 76Canc competed with 6F5 and targeted the 6HB similarly to 6F5 and 6F11, we then assessed its ADCC function. 76Canc was surprisingly comparable in activity to 6F5 and 6F11 against both clad B and C targets (**Figure 6**). despite having relatively lower EC50 (**Figure 4**) and no mutations away from predicted germline heavy chain outside of the CDR3 region (**Figure 3**).

**Figure 6:**
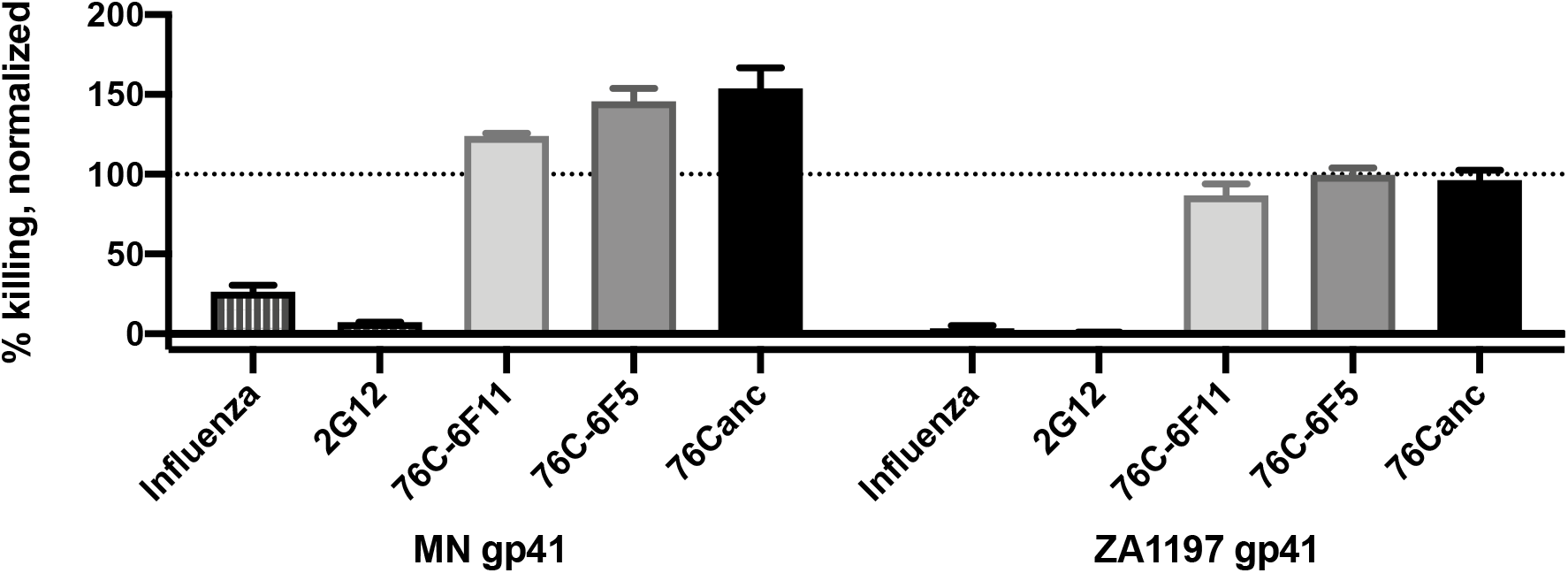
76Canc, with germline variable segment VH1-02 containing Ab from group 76C, retains ADCC activity. ADCC activity of 76C-6F11, 76C-6F5, and 76Canc using clade C ZA1197 gp41 and clade B MN gp41. Data are normalized to HIV immunoglobulin (HIVIG; represented by dotted line) with influenza-specific MAb Ab F16V3 and HIV bnAb 2G12 shown as controls. Results were repeated twice with representative data of single experiment shown.

## Discussion

Elicitation of broadly neutralizing Abs remains a goal of HIV vaccine research. While these Abs can arise during HIV-1 infection, our limited knowledge of their germline, pre-immune Ab responses limits our ability to elicit them through vaccination (Yacoob et al., 2016). We show here that an Ab with germline VH1-02 can recognize and show appreciable ADCC activity against HIV. This ADCC activity with germline variable segment contributions, the diversity of these mature clones, and the initial cross-reactive binding studies shown here support that these Abs were formed from naïve B cell responses. This finding has direct implications on current vaccine regimens. Identification of epitopes that can be recognized by germline antibodies with functional activity may allow for a more direct route for vaccination (single antigen and/or low number of booster approach) (Stamatatos, Pancera, & McGuire, 2017) rather than complicated multiple boosting regimens with variable antigens (Saunders et al., 2019).

Highly mutated anti-CD4bs bnAbs preferentially use the VH1 gene, in particular the VH 1-02 and 1-46 segments, as the heavy chain predominantly drives CD4 binding site recognition (Scheid et al., 2011; Wu et al., 2010). VRC01 is highly mutated, with V-gene region amino acid predicted mutations of 42% in the VH1-02 heavy chain and 28% in the light chain (Wu et al., 2010; Zhou et al., 2010). There is structural convergence of VRC01 related Abs and these rely on short CDRs in chosen light chains (Zhou et al., 2010; Zhou et al., 2013). In contrast to 76Canc, unmutated common ancestors of VRC01 Abs don‘t interact with native trimers(Hoot et al., 2013). This potentially could interfere with vaccination strategies based on the concept of stimulating the naïve Ab to generate a HIV bnAb response(Hoot et al., 2013; McGuire et al., 2013; Zhou et al., 2010). Select framework mutations in VRC01 to ‘near ‘ germline did maintain functionality (Georgiev et al., 2014). Recently, recombinant humanized immunoglobulin mouse models using VH1-02 sequences and a sequential immunization strategy have shown success in creating similarly matured Abs (Tian et al., 2016). Potentially these strategies can be utilized to similarly target 76C-like Abs.

As the members of 76C showed general structural homology, but few shared mutations, 76C-like Abs may arise from a truly naïve initial response. However, Abs with low numbers of mutations may have been originally focused on other antigens. A number of the HIV bnAbs have been described as having autoimmune potential (Zandman-Goddard & Shoenfeld, 2002). As prior studies have suggested, gp41 targeting during initial infection relies predominantly on stimulating memory B cells that have previously been activated by non–HIV-1 antigens. A similar study with germline reversion of gp41 targeting Abs has been performed; however, germline Abs lacked HIV reactivity (Liao et al., 2011). These Abs were found to be poly-reactive and bound to various host or gut flora antigens. A number of gut ileum B cell derived anti-gp41 Abs cross-reacted to commensal bacteria and showed non-HIV-1 polyreactivity (Trama et al.). Groups have postulated that germline Abs primed by reactions to commensal bacteria can be stimulated and form the basis for anti-gp41 Ab responses after infection.

The original antigen that was recognized by these clones remains unknown. We show that the 76Canc binds the 6HB comparable to other 76C Abs, but is less efficient at competing against 6F5. It may be that a structure similar to the 6HB was the original selective antigen. Since these Abs were cloned from LTNPs and similar Abs are more prevalent in LTNPs, it is intriguing to consider that the potential effects of this type of functional Ab response early in an infection could influence the global clinical course of disease.

### Conclusions

Here we show that the VH1-02 germline variable chain can support ADCC. Exploring the targeting of ancestor Abs such as this may help inform on future vaccine efforts. The shepherding of a naïve response all the way to highly mutated Abs remains a challenge. Targeting efforts to expand functional Abs utilizing germline sequences, such as 76Canc, may offer a simpler path to immune protection or assist in immunity during vaccination with more complex regimens.

## Data Availability

All pertinent data is included in the manuscript. Raw data is available on request.

## Acknowledgements

We thank Dr. Julie Overbaugh for her comments on previous versions of this manuscript. We thank Dr. J Bloom for providing influenza-specific monoclonal Ab F16V3. The following reagents were obtained through the AIDS Research and Reference Reagent Program, Division of AIDS, NIAID, NIH: monoclonal Abs 2G12 from Dr. Hermann Katinger; anti-HIV-1 gp41 monoclonal Ab NC-1 from Dr. Shibo Jiang. This research was supported through biorepository access by Research reported in this [publication/press release] was supported by the University of Washington / Fred Hutch Center for AIDS Research, an NIH-funded program under award number AI027757 which is supported by the following NIH Institutes and Centers: NIAID, NCI, NIMH, NIDA, NICHD, NHLBI, NIA, NIGMS, NIDDK. Study was supported by NIH R01 AI 125119-01; “The role of non-broadly neutralizing antibodies targeting gp41 structural epitopes in long term nonprogression of HIV infection.”

## References

Bonsignori, M., Pollara, J., Moody, M. A., Alpert, M. D., Chen, X., Hwang, K. K., … Haynes, B. F. (2012). Antibody-dependent cellular cytotoxicity-mediating antibodies from an HIV-1 vaccine efficacy trial target multiple epitopes and preferentially use the VH1 gene family. J Virol, 86(21), 11521–11532. doi:10.1128/JVI.01023-12

Burton, D. R., Hessell, A. J., Keele, B. F., Klasse, P. J., Ketas, T. A., Moldt, B., … Moore, J. P. (2011). Limited or no protection by weakly or nonneutralizing antibodies against vaginal SHIV challenge of macaques compared with a strongly neutralizing antibody. Proc Natl Acad Sci U S A, 108(27), 11181–11186. doi:10.1073/pnas.1103012108

Das, J., Devadhasan, A., Linde, C., Broge, T., Sassic, J., Mangano, M., … Alter, G. (2020). Mining for humoral correlates of HIV control and latent reservoir size. PLoS Pathog, 16(10), e1008868. doi:10.1371/journal.ppat.1008868

Georgiev, I. S., Rudicell, R. S., Saunders, K. O., Shi, W., Kirys, T., McKee, K., … Kwong, P. D. (2014). Antibodies VRC01 and 10E8 neutralize HIV-1 with high breadth and potency even with Ig-framework regions substantially reverted to germline. J Immunol, 192(3), 1100–1106. doi:10.4049/jimmunol.1302515

Haynes, B. F., Gilbert, P. B., McElrath, M. J., Zolla-Pazner, S., Tomaras, G. D., Alam, S. M., … Kim, J. H. (2012). Immune-correlates analysis of an HIV-1 vaccine efficacy trial. N Engl J Med, 366(14), 1275–1286. doi:10.1056/NEJMoa1113425

Hessell, A. J., Hangartner, L., Hunter, M., Havenith, C. E., Beurskens, F. J., Bakker, J. M., … Burton, D. R. (2007). Fc receptor but not complement binding is important in antibody protection against HIV. Nature, 449(7158), 101–104. doi:10.1038/nature06106

Hessell, A. J., Rakasz, E. G., Tehrani, D. M., Huber, M., Weisgrau, K. L., Landucci, G., … Burton, D. R. (2009). Broadly Neutralizing Monoclonal Antibodies 2F5 and 4E10, Directed Against the Human Immunodeficiency Virus Type 1 (HIV-1) gp41 Membrane Proximal External Region (MPER), Protect Against SHIVBa-L Mucosal Challenge. J Virol. doi:JVI.01272-09 [pii] 10.1128/JVI.01272-09

Hicar, M. D., Chen, X., Kalams, S. A., Sojar, H., Landucci, G., Forthal, D. N., … Crowe, J. E., Jr. (2016). Low frequency of broadly neutralizing HIV antibodies during chronic infection even in quaternary epitope targeting antibodies containing large numbers of somatic mutations. Mol Immunol, 70, 94–103. doi:10.1016/j.molimm.2015.12.002

Hicar, M. D., Chen, X., Sulli, C., Barnes, T., Goodman, J., Sojar, H., … Crowe, J. E., Jr. (2016). Human Antibodies that Recognize Novel Immunodominant Quaternary Epitopes on the HIV-1 Env Protein. PLoS One, 11(7), e0158861. doi:10.1371/journal.pone.0158861

Hoot, S., McGuire, A. T., Cohen, K. W., Strong, R. K., Hangartner, L., Klein, F., … Stamatatos, L. (2013). Recombinant HIV envelope proteins fail to engage germline versions of anti-CD4bs bNAbs. PLoS Pathog, 9(1), e1003106. doi:10.1371/journal.ppat.1003106

Jiang, S., Lin, K., & Lu, M. (1998). A conformation-specific monoclonal antibody reacting with fusion-active gp41 from the human immunodeficiency virus type 1 envelope glycoprotein. J Virol, 72(12), 10213–10217.

Lambotte, O., Ferrari, G., Moog, C., Yates, N. L., Liao, H. X., Parks, R. J., … Delfraissy, J. F. (2009). Heterogeneous neutralizing antibody and antibody-dependent cell cytotoxicity responses in HIV-1 elite controllers. AIDS, 23(8), 897–906. doi:10.1097/QAD.0b013e328329f97d

Liao, H. X., Chen, X., Munshaw, S., Zhang, R., Marshall, D. J., Vandergrift, N., … Haynes, B. F. (2011). Initial antibodies binding to HIV-1 gp41 in acutely infected subjects are polyreactive and highly mutated. J Exp Med, 208(11), 2237–2249. doi:10.1084/jem.20110363

Liu, S., Zhao, Q., & Jiang, S. (2003). Determination of the HIV-1 gp41 fusogenic core conformation modeled by synthetic peptides: applicable for identification of HIV-1 fusion inhibitors. Peptides, 24(9), 1303–1313. doi:10.1016/j.peptides.2003.07.013

McGuire, A. T., Hoot, S., Dreyer, A. M., Lippy, A., Stuart, A., Cohen, K. W., … Stamatatos, L. (2013). Engineering HIV envelope protein to activate germline B cell receptors of broadly neutralizing anti-CD4 binding site antibodies. J Exp Med, 210(4), 655–663. doi:10.1084/jem.20122824

Milligan, C., Richardson, B. A., John-Stewart, G., Nduati, R., & Overbaugh, J. (2015). Passively acquired antibody-dependent cellular cytotoxicity (ADCC) activity in HIV-infected infants is associated with reduced mortality. Cell Host Microbe, 17(4), 500–506. doi:10.1016/j.chom.2015.03.002

Parren, P. W., Ditzel, H. J., Gulizia, R. J., Binley, J. M., Barbas, C. F., 3rd, Burton, D. R., & Mosier, D. E. (1995). Protection against HIV-1 infection in hu-PBL-SCID mice by passive immunization with a neutralizing human monoclonal antibody against the gp120 CD4-binding site. AIDS, 9(6), F1–6.

Parren, P. W., Fisicaro, P., Labrijn, A. F., Binley, J. M., Yang, W. P., Ditzel, H. J., … Burton, D. R. (1996). In vitro antigen challenge of human antibody libraries for vaccine evaluation: the human immunodeficiency virus type 1 envelope. J Virol, 70(12), 9046–9050.

Santra, S., Tomaras, G. D., Warrier, R., Nicely, N. I., Liao, H. X., Pollara, J., … Haynes, B. F. (2015). Human Non-neutralizing HIV-1 Envelope Monoclonal Antibodies Limit the Number of Founder Viruses during SHIV Mucosal Infection in Rhesus Macaques. PLoS Pathog, 11(8), e1005042. doi:10.1371/journal.ppat.1005042

Saunders, K. O., Wiehe, K., Tian, M., Acharya, P., Bradley, T., Alam, S. M., … Haynes, B. F. (2019). Targeted selection of HIV-specific antibody mutations by engineering B cell maturation. Science, 366(6470). doi:10.1126/science.aay7199

Scheid, J. F., Mouquet, H., Ueberheide, B., Diskin, R., Klein, F., Oliveira, T. Y., … Nussenzweig, M. C. (2011). Sequence and structural convergence of broad and potent HIV antibodies that mimic CD4 binding. Science, 333(6049), 1633–1637. doi:10.1126/science.1207227

Smith, M., Hoffman, J., Sojar, H., Aalinkeel, R., Hsiao, C. B., & Hicar, M. D. (2018). Assessment of Antibody Interference of Enfuvirtide (T20) Function Shows Assay Dependent Variability. Curr HIV Res, 16(6), 404–415. doi:10.2174/1570162X17666190228154850

Sojar, H., Baron, S., Sullivan, J. T., Garrett, M., van Haaren, M. M., Hoffman, J., … Hicar, M. D. (2019). Monoclonal Antibody 2C6 Targets a Cross-Clade Conformational Epitope in gp41 with Highly Active Antibody-Dependent Cell Cytotoxicity. J Virol, 93(17). doi:10.1128/JVI.00772-19

Stamatatos, L., Pancera, M., & McGuire, A. T. (2017). Germline-targeting immunogens. Immunol Rev, 275(1), 203–216. doi:10.1111/imr.12483

Tian, M., Cheng, C., Chen, X., Duan, H., Cheng, H. L., Dao, M., … Alt, F. W. (2016). Induction of HIV Neutralizing Antibody Lineages in Mice with Diverse Precursor Repertoires. Cell, 166(6), 1471–1484 e1418. doi:10.1016/j.cell.2016.07.029

Trama, Ashley M., Moody, M. Anthony, Alam, S. Munir, Jaeger Frederick H., Lockwood, Bradley, Parks, Robert, … Haynes Barton F. (2014). HIV-1 envelope gp41 antibodies can originate from terminal ileum B cells that share cross-reactivity with commensal bacteria. Cell host & microbe, 16(2), 215–226.

Williams, K. L., Cortez, V., Dingens, A. S., Gach, J. S., Rainwater, S., Weis, J. F., … Overbaugh, J. (2015). HIV-specific CD4-induced Antibodies Mediate Broad and Potent Antibody-dependent Cellular Cytotoxicity Activity and Are Commonly Detected in Plasma From HIV-infected humans. EBioMedicine, 2(10), 1464–1477. doi:10.1016/j.ebiom.2015.09.001

Wu, X., Yang, Z. Y., Li, Y., Hogerkorp, C. M., Schief, W. R., Seaman, M. S., … Mascola, J. R. (2010). Rational design of envelope identifies broadly neutralizing human monoclonal antibodies to HIV-1. Science, 329(5993), 856–861. doi:10.1126/science.1187659

Yacoob, C., Pancera, M., Vigdorovich, V., Oliver, B. G., Glenn, J. A., Feng, J., … Stamatatos, L. (2016). Differences in Allelic Frequency and CDRH3 Region Limit the Engagement of HIV Env Immunogens by Putative VRC01 Neutralizing Antibody Precursors. Cell Rep, 17(6), 1560–1570. doi:10.1016/j.celrep.2016.10.017

Zandman-Goddard, G., & Shoenfeld, Y. (2002). HIV and autoimmunity. Autoimmun Rev, 1(6), 329–337. doi:10.1016/s1568-9972(02)00086-1

Zhou, T., Georgiev, I., Wu, X., Yang, Z. Y., Dai, K., Finzi, A., … Kwong, P. D. (2010). Structural basis for broad and potent neutralization of HIV-1 by antibody VRC01. Science, 329(5993), 811–817. doi:10.1126/science.1192819

Zhou, T., Zhu, J., Wu, X., Moquin, S., Zhang, B., Acharya, P., … Kwong, P. D. (2013). Multidonor analysis reveals structural elements, genetic determinants, and maturation pathway for HIV-1 neutralization by VRC01-class antibodies. Immunity, 39(2), 245–258. doi:10.1016/j.immuni.2013.04.012

